# Circannual prevalence of vitamin D insufficiency in older and minoritized ethnic adults in Northern Britain: screening outcomes from a clinical trial (ISRCTN13778806)

**DOI:** 10.1101/2025.11.17.25340416

**Authors:** Alice Goddard, Anthony Watson, Rowena Tilbury, Bernard M. Corfe, Andrea Fairley

## Abstract

Vitamin D is essential for bone and metabolic health. Deficiency remains a global health issue, particularly among older adults and ethnic minorities with darker skin pigmentation. Data on circannual variation these groups remain sparse.

This study reports vitamin D status in older adults (≥65 years) and ethnic adults (≥18 years, Fitzpatrick classes IV–VI) in northern Britain during the screening phase of a supplementation trial. Participants were screened for inclusion between December 2024 and August 2025. Serum 25-hydroxyvitamin D (25(OH)D) was assessed in dried blood spots followed by LC-MS/MS analysis.

299 participants were screened. Vitamin D insufficiency or deficiency (<50 nmol/L) was noted in 54.8% of older adults and 72.1% of ethnic individuals. These rates did not decline during summer months. These findings highlight persistently high rates of vitamin D insufficiency across high-risk groups in northern Britain and underscore the inadequacy of sunlight exposure as a corrective measure.

## Introduction

Vitamin D is a fat-soluble vitamin essential for bone development and maintenance, primarily by enhancing calcium, magnesium, and phosphate absorption. It is obtained through dietary sources, mainly as cholecalciferol (vitamin-D_3_) found in animal-based foods and as ergocalciferol (vitamin-D_2_) derived from plant sources and UV-exposed fungi. Additionally, dermal exposure to sunlight (ultraviolet-B) enables cutaneous synthesis of vitamin D through the photoconversion of 7-dehydrocholesterol to previtamin-D_3_ and thermal isomerisation to form cholecalciferol.

Vitamin D is absorbed in the small intestine and transported to the liver by chylomicrons before undergoing both hepatic and renal hydroxylation, forming key metabolites, 25-hydroxyvitaminD (25(OH)D), the primary circulating form, and 1,25-dihydroxyvitaminD (1,25(OH)2D), the active hormonal form, respectively. Whilst both are necessary for biological functions, 25(OH)D, is the most widely recognised as a reliable biomarker for assessing vitamin D status in humans [1]. Sufficient levels are defined as >50 nmol/L, with insufficiency at 31-49 nmol/L and deficiency at <30 nmol/L [2]. Supplementation, achieved through specific food supplements or fortified dietary sources, is recommended to maintain adequate vitamin D levels in various high-risk populations [3].

Vitamin D deficiency is a widespread public health concern linked not only to bone and immune health, but also to increased risk of chronic diseases such as cardiovascular disease and diabetes [4-5]. Although vitamin D deficiency is commonly viewed as a seasonal issue due to reduced sunlight exposure during winter, emerging evidence suggests that some demographic groups remain at risk year-round. Within the UK, certain subpopulations including various ethnic groups are at greater risk of vitamin D deficiency. In 2021, up to 92% of UK-dwelling South Asians (n = 5918, aged 40-69 years and 84% of UK-dwelling African-Caribbean individuals (n = 4046, aged ≥40 years) had insufficient 25(OH)D levels (<50 nmol/L) [6-7] demonstrating how individuals within these communities are at greater risk of deficiency. Likewise, deficiency is widespread among older adults globally, occurring irrespective of season [8] reinforcing the need for targeted public health strategies addressing these vulnerable groups.

Our current study (ISRCTN13778806) investigates the nutrikinetics of vitamin D supplementation in older adults and ethnic individuals with darker skin pigmentation (as quantified by the Fitzpatrick classes IV, V, VI) currently residing in Britain. This report focuses on the screening phase of the study, providing an overview of baseline demographic characteristics and the prevalence of vitamin D deficiency and insufficiency among people screened for the trial.

## Methods

The trial strata were older adults (65 years and above) and ethnic adults aged 18 years and above with a darker skin pigmentation, Fitzpatrick classes IV, V, VI. The inclusion criteria encompassed individuals currently residing in Britain, including those aged 65 years or older of any ethnic background, as well as individuals aged 18 years or older with mixed, multiple, or minority ethnic heritage. All participants were screened for their vitamin D status and only those who had a suboptimal vitamin D status (25(OH)D < 50 nmol/L) were enrolled onto the main trial.

Participants were recruited using a multimodal strategy combining digital, community based and in-person approaches. Eligible participants attend an appointment for anthropometric measurements and self-administered finger-prick dried blood spot (DBS) test to assess baseline 25(OH)D levels. The DBS cards were posted to and analysed independently by SureScreen Scientifics Ltd (Morley Retreat, Church Lane, Morley, Derbyshire, DE7 6DE) using an LC-MS/MS analytical protocol [9].

## Results

Recruitment opened in December 2024 and closed in August 2025. In total, 936 candidates approached the study, with 630 candidates being excluded due to ineligibility. The most common exclusion criteria were already taking cholecalciferol supplementation and withdrawal due to disinterest or lifestyle suitability. In total 299 individuals were screened and their data were included in this analysis. The CONSORT is shown in **Fig 1A**. Participant demographics are shown in **Table 1**.

**Table 1.**
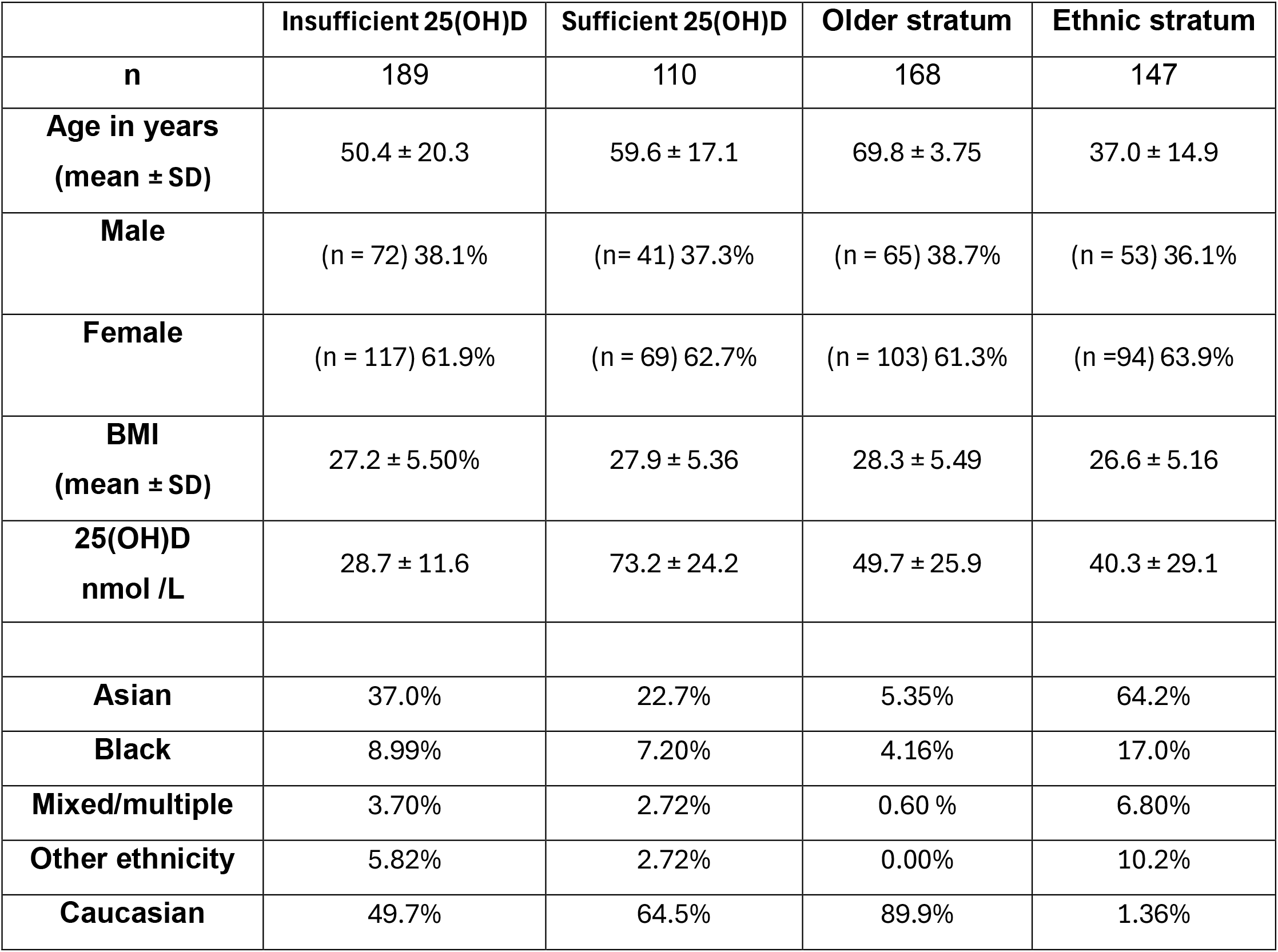
Summary of demographic profile of the study population.

**Figure 1.**
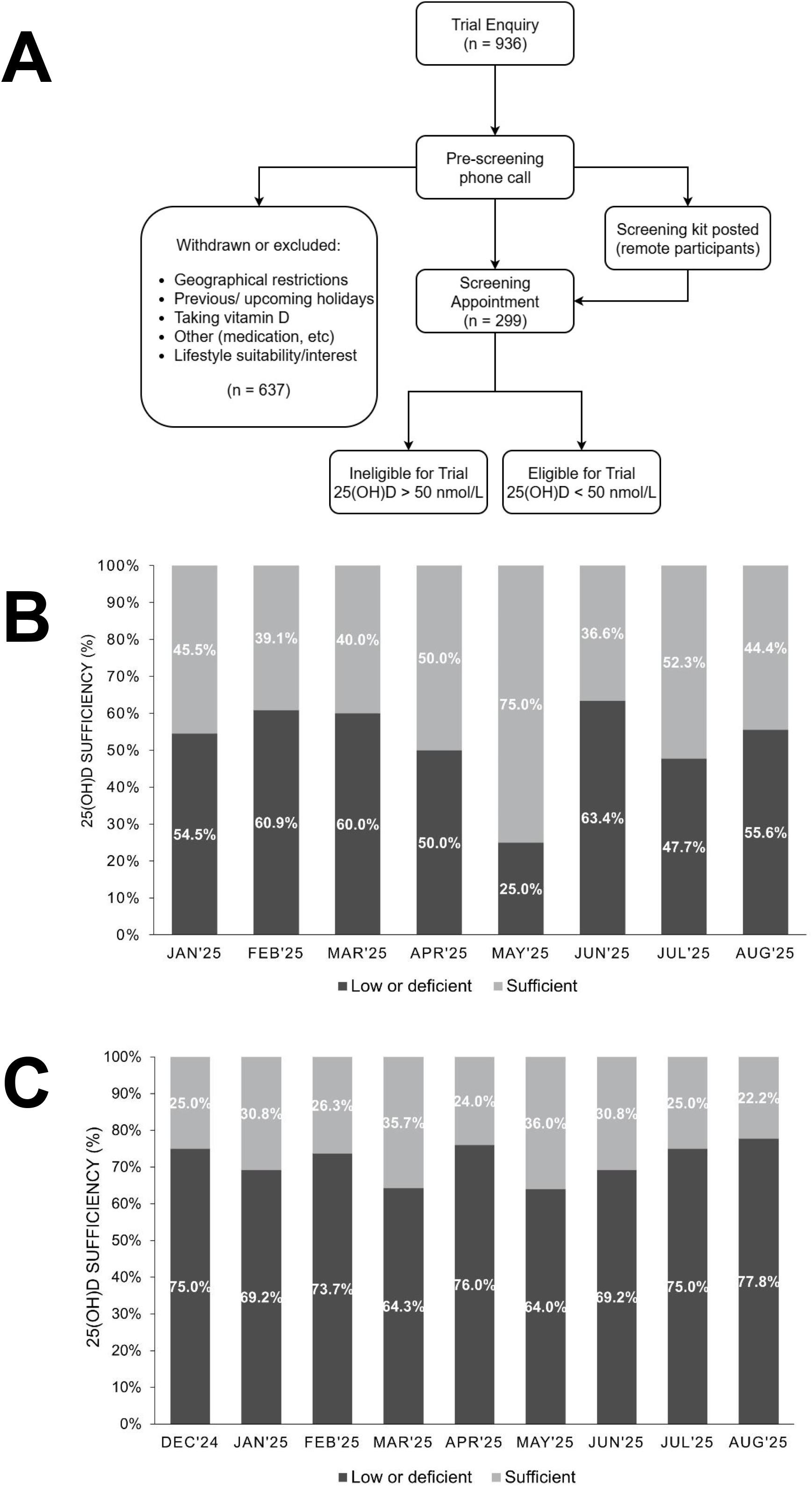
Summary of key study outcomes. **Panel A** shows the CONSORT for participants included in this study. **Panel B** shows the prevalence of vitamin D insuficiency and deficiency by month of recruitment in the older adult stratum of the of the study. **Panel C** shows the prevalence of vitamin D insuficiency and deficiency by month of recruitment in the ethnic adults stratum of the of the study.

Prevalence of low 25(OH)D status was assessed month by both across screening dates (December-August 2025). Among all older adults assessed (≥65 years, n = 168), 54.8% were classified as insufficient 25(OH)D (low or deficient) there was little variation by trial reporting month (**Fig 1B**). In contrast, ethnic individuals (≥18 years, n = 147), showed consistently higher rates of 25(OH)D insufficiency, with 72.1% classified as low or deficient with no changes in the proportion of individuals who were insufficient across seasons, winter, spring, and summer (**Fig 1C**). Recruitment targets were met in August, so no additional data was collected thereon.

The prevalence of vitamin D insufficiency within each trial strata, and further subanalyses by ethnic subgroup can be found in **Supplementary Figure S1**.

## Discussion

Our study reveals a consistently high and potentially prolonged prevalence of vitamin D insufficiency among Britain’s older and ethnic individuals with monthly 25(OH)D deficiency prevalence rates not falling below 25.0% for older adults and 64.0% for ethnic minorities. Despite the assumption that summer sunlight improves vitamin D status, our data shows that older adults continued to experience relatively high deficiency rates even during summer with an average insufficiency of 55.6% (June-August). This substantially exceeds the approximate 20% year-round deficiency rate observed in the general population [10]

More than half of older adults remained deficient even in periods of peak sunlight, indicating that seasonal variation may have limited impact in mitigating insufficiency. The trend was even more pronounced among ethnic individuals. In this strata, deficiency is likely attributed to broader environmental and or lifestyle factors, such as diet, skin pigmentation, and cultural practices [11].

These findings challenge the reliance on sunlight exposure as the primary strategy for repleting suboptimal 25(OH)D status during summer months in northern britain. As such, current guidelines on seasonal adjustments to vitamin D intake may not be appropriate for these populations, especially in northern latitudes. It is crucial to consider year-round supplementation and implement more targeted public health strategies [12]. Interventions should address specific dietary needs and cultural barriers, promoting a more comprehensive approach to tackling vitamin D insufficiency across high-risk groups.

## Funding

This project was wholly funded by BetterYou Ltd

## Compliance with Ethical Standards

### Conflict of Interest

BetterYou funded this study. This sponsor was not involved in the study design, delivery or interpretation of the data, which was undertaken entirely by Newcastle University. The authors declare that they have no conflict of interest

## Data availability

Corfe, Bernard (2025). Dataset for screening phase of a clinical trial ISRCTN13778806. figshare. Dataset. https://doi.org/10.6084/m9.figshare.30636023.v1

## Supplementary analyses

**Figure S1.**
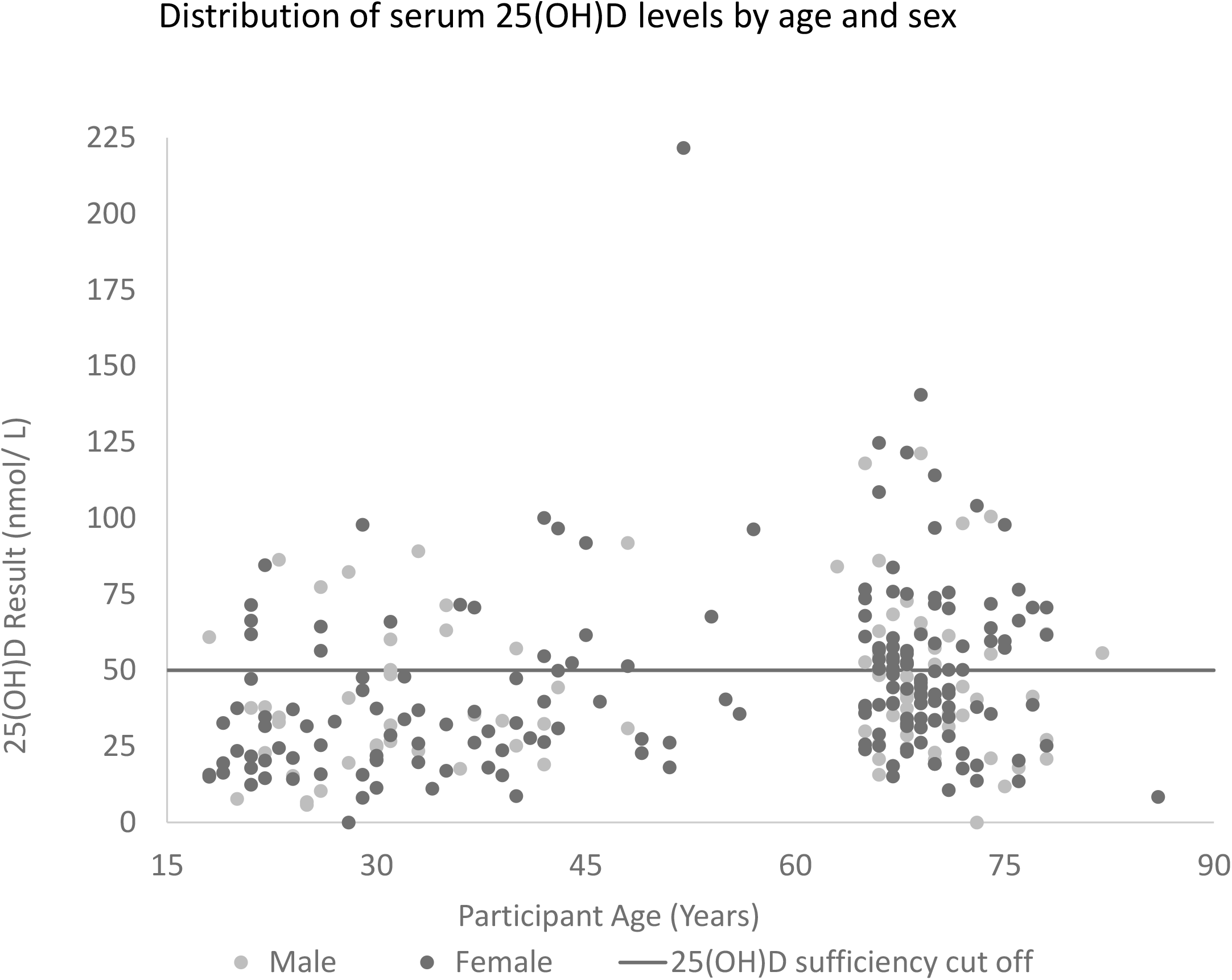
Distribution of serum 25(OH)D levels by age and sex for 299 screened participants in relation to vitamin D insufficiency, below 50 nmol/ L.

**Figure S2.**
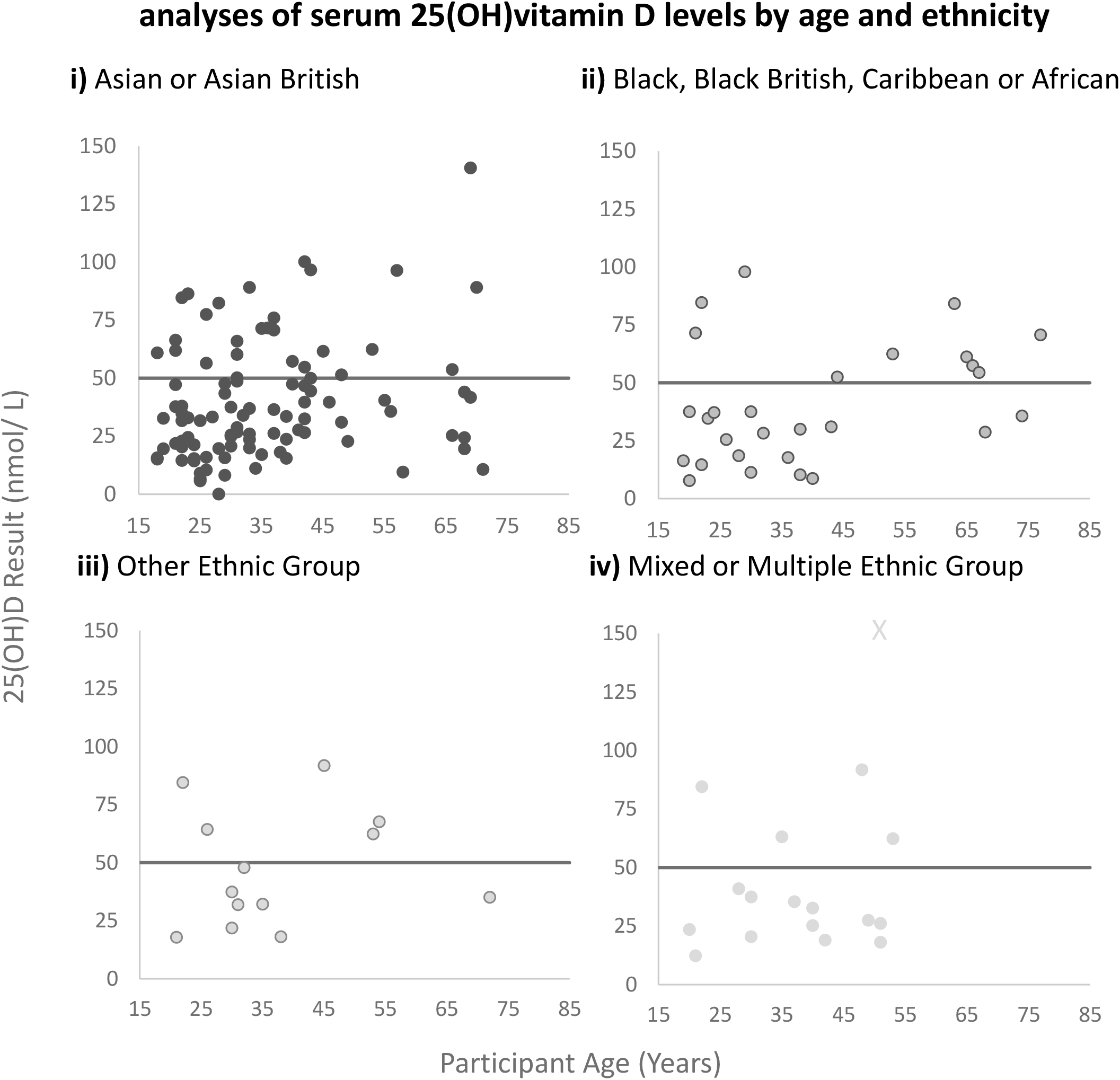
analyses of serum 25(OH)vitamin D levels by age and ethnicity. Distribution of serum 25(OH)D levels by age within the ethnic adult stratum **i)** Asian or Asian British, **ii)** Black, Black British, Caribbean or African, **iii)** Mixed or multiple ethnic groups, and **iv)** Other ethnic group.

## Notes

### Competing Interest Statement

The authors have declared no competing interest.

### Clinical Trial

ISRCTN13778806

### Author Declarations

Faculty of Medical Sciences Ethical Committee, Newcastle University gave ethical approval for this work

